# Comparison of three nasopharyngeal swab types and the impact of physiochemical properties for optimal SARS-CoV-2 detection

**DOI:** 10.1101/2020.10.21.20206078

**Authors:** Trish R. Kahamba, Lara Noble, Wendy Stevens, Lesley Scott

## Abstract

Adequate swab specimen collection, release and detection of nucleic acids by molecular diagnostic assays is largely attributed to the physical and chemical characteristics of different swab types. We investigated properties of three types of commercial nasopharyngeal swabs (nylon flocked: Type 1-Media Merge; Type 2-Kang Jian Medical Apparatus, China and Type 3-Wuxi NEST Biotechnology Co. Ltd, China) used in clinical diagnostics with the aim to establish if different swab designs and configurations had any effect on swab performance. Properties investigated included viral absorption, release, capture, extraction and recovery efficiency from each swab for the detection of Severe Acute Respiratory Syndrome Coronavirus-2 (SARS-CoV-2). All swab types (n=18) were inoculated with different amounts of SARS-CoV-2 live viral cultures (1:10, 1:100 and 1:1000 copies/ml) and eluted in sterile phosphate buffer saline. RNA was extracted from all swab eluates using a fully automated system (BD MAX™ System) and cycle threshold (Ct) values were compared. RNA stability was also investigated after dry storage of swabs at room temperature for 72 hours. Statistically significant differences (p<0.05) were observed in the absorption and release capabilities between Type 1 and 3 as well as between Type 2 and 3 swabs, however, no significant difference was observed between Type 1 and 2. Ct values and extraction efficiency amounts of SARS-CoV-2 varied amongst the swab types. We conclude that in order to facilitate accurate SARS-CoV-2 diagnosis, assessment of NP swab characteristics is of importance before implementation for specimen collection in the clinical setting.

## Introduction

Accurate diagnosis of respiratory infections using swabs as the specimen collection device is largely dependent on the types of fibres and their physical characteristics (1, 2). Moreover, the sampling techniques i.e. pressure applied, swab rotations, number of strokes and sampling site are paramount for sufficient organism capture and diagnostic sensitivity (3). As recommended by the World Health Organization (WHO) and US Centers for Disease Control and Prevention (CDC), the use of the nasopharyngeal (NP) swab is the main sampling method to detect Severe Acute Respiratory Syndrome Coronavirus-2 (SARS-CoV-2) (4, 5). It is further recommended that sampling of the secretions from the posterior nasopharynx must be conducted using only synthetic fiber swabs like rayon or nylon flocked swabs with plastic or wire shafts (4).

Both nylon flocked and rayon swabs have been shown to have similar efficiency in preserving influenza RNA (6). Conversely, a study by Daley *et al*., have shown better yields of respiratory epithelial cells by flocked than rayon swabs after NP sampling (7). Hernes *et al*., also reported that nylon flocked swabs are more efficient than rayon swabs (8).

Studies focusing on SARS-CoV-2 have demonstrated that NP swabs are more sensitive than oropharyngeal (OP) swabs (9, 10), however, there are other studies that have shown comparable sensitivities between the two swab types (11). It has been previously reported that the quality of swab samples contribute to proper disease detection and monitoring of respiratory infections such as avian influenza (12) and swab composition and structure significantly impact the collection and release efficiency of a sample (13, 14).

During the period of June 2020, our laboratory at the Department of Molecular Medicine and Haematology at University of Witwatersrand received NP swabs from three different manufacturers for use in SARS-CoV-2 research. We noticed very distinct differences of the designs and sizes of the swab tip as well as the amount of nylon flock material on the swabs. We then aimed to investigate the efficiency of the three NP swabs in terms of viral capture, absorption, extraction efficiency as well as to compare the qualitative characteristics i.e. material type, length of shaft, breakpoint, ease of use and robustness. PCR inhibition was also monitored for all three NP swabs.

## Materials and Methods

### Swabs

We investigated three types of NP (nylon flocked) swabs from different manufacturers: Wuxi NEST Biotechnology Co. Ltd, China, Kang Jian Medical Apparatus, China and Media Merge (South African supplier-manufacturer undisclosed). All swabs had plastic shafts and breakpoints and were supplied individually wrapped in sterile plastic pouches. (Figure 1). Swab characteristics such as shaft length, breakpoint, ease of use and robustness were assessed using a 5-point Likert scale.

**Figure 1:**
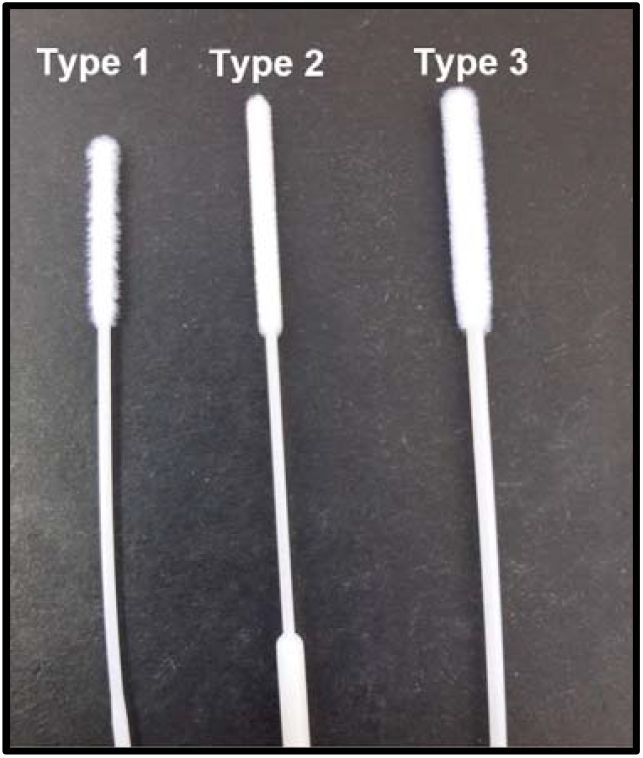
Nasopharyngeal nylon flocked swabs investigated. Type 1-South African supplier; Type 2- Kang Jian Medical Apparatus, China and Type 3- Wuxi NEST Biotechnology Co. Ltd, China.

### Swab volume absorption and release capacity

In order to determine the absorption capacity and volume released of each swab, we adopted methods and formulae for calculations from Zasada *et al*., and Wanke *et al*., (14, 15) with some modifications. Briefly, swabs were immersed for 5-10s in pre-weighed 1.5ml eppendorf tubes that contained pre-aliquotted spiked phosphate buffered saline (PBS) (500µl) (Diagnostics Media Products, National Health Laboratory Service, South Africa) spiked with SARS-CoV-2 live viral cultures. The swabs were removed and the eppendorf tubes were weighed again. The weight and volume of liquid absorbed as well as volume released were calculated using the following formulae:

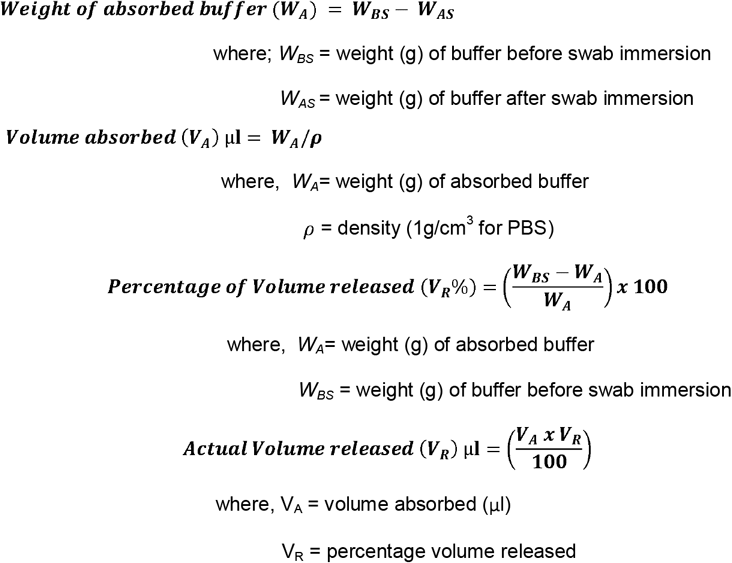

### Swab viral capture, extraction and recovery efficiency analysis

Swab capture was calculated using information derived from Moore *et al*., (16) with some modifications to the formula, i.e. for this study we inferred capture to be the same as removal of viral particles from buffer hence we used the following formula:

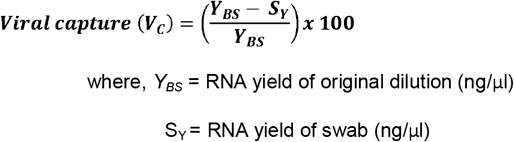

Swab extraction efficiency of RNA was determined using modified methods and formula from Bruijins *et al*., (17) and Zasada *et al*., (14). Briefly swabs were spiked with viral cultures, followed by elution in 1.5ml PBS. RNA was extracted from the swab eluates (750µl) using the BD SARS-CoV-2 reagents on the BD MAX™ System (Becton, Dickinson and Company, Maryland, USA) according to manufacturer instructions (18). Extracted RNA was quantified using the NanoDrop™ 2000 Spectrophotometer (Thermo Fisher Scientific™ Inc., USA) and the extraction efficiency of each swab was calculated using the following formula:

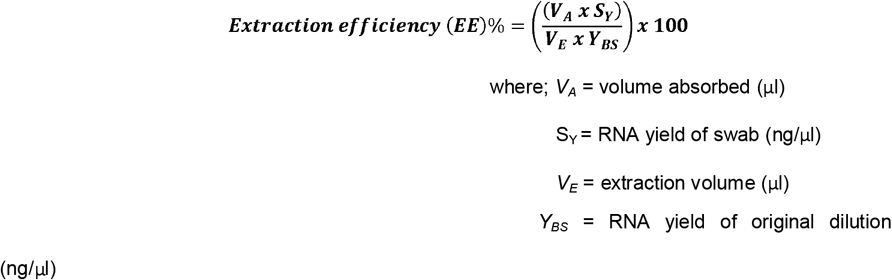

Recovery efficiency was derived from information from Rose *et al*., (19) and calculated using the following formula:

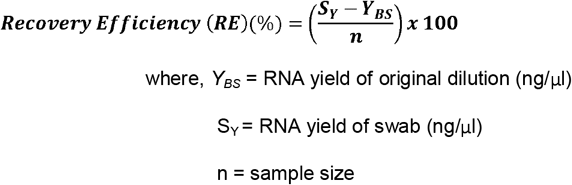

### Preparation of viral cultures and inoculation of swabs

SARS-CoV-2 live viral cultures obtained from the Centre of Excellence for Biomedical TB Research at the University of the Witwatersrand were spiked into PBS and then serially diluted to final concentrations of 1:10, 1:100, and 1:1000 copies/ml. All swab types were inserted into each dilution (5-10s with swirling), transferred into fresh 1.5ml PBS followed by a vortex step for 5-10s. Positive and negative controls were included in both assays. We tested the PBS before spiking as the negative control and positive controls were derived from the highest concentrations i.e. 1:10 copies/ml SARS-CoV-2. Eluates were stored at 2-8°C and tested within 1 hour (day 0).

### RNA stability on different swabs

Some of the inoculated swabs were transferred into dry 1.5ml eppendorf tubes and stored for 72 hours (day 3) in zip-lock bags at room temperature in order to simulate transportation from sampling location to the laboratory after which swabs were resuspended in 1.5ml PBS, vortexed 5-10s and stored at 2-8°C. Testing was performed within 1hour.

### Swab Testing: real-time RT-PCR

Swab eluates from the different dilutions were extracted using real-time RT-PCR assay (BD SARS-CoV-2 reagents) on the BD MAX™ System according to manufacturer instructions (18). Testing was performed at two time points-day 0 and day 3. Cycle threshold (Ct) values of each swab eluate were compared.

### Statistical analysis

We performed the ANOVA statistical test using Stata/SE 16.0 (StataCorp LLC, USA) to determine differences among means. When statistical significant differences were indicated (p<0.05), post-hoc comparison was performed using pairwise methods (sidak, bonferroni and scheffe) to determine the means that were significantly different from each other.

## Results

### Qualitative assessments

Scores using a 5-point Likert scale were given by the user of the swabs. Swab material was scored based on texture, odor and color of swab tip. Type 1 swab had the softest brush-like texture and was given a score of 5. All swabs were odorless. Type 1 and type 3 swab tips appeared to be the whitest in color (indicating high purity) and were therefore given a score of five. All shafts were made of plastic, hence all swabs scored 5. Shaft properties included flexibility, brittleness and debris production when broken beyond the breakpoint. All swab types were tough and did not produce any debris when broken, however, types 1 and 2 appeared more flexible compared to type 3. Type 1 was the shortest in length and types 2 and 3 were almost the same lengths. All swabs had good breakpoints, but type 2 could not be snapped off and the breakpoint dismantled after a few twists. All swabs were easy to use and did not disintegrate after vortexing/centrifugation (3000rpm, 5mins). Overall swab types 1 and 2 were given the highest scores. (Refer to supplementary table S1 for more information).

### Swab volume absorption and release capacity

The buffer absorption capacity of the three swab types ranged from 55.5 to 125.8 µL. Swab type 3 exhibited the highest absorption capacity compared to the remaining two types. The ability to absorb and release buffer of swab type 3 was statistically greater than types 1 and 2; (*p*<0.05). Differences in absorption of types 1 and 2 were minimal and not statistically different (Figure 2).

**Figure 2:**
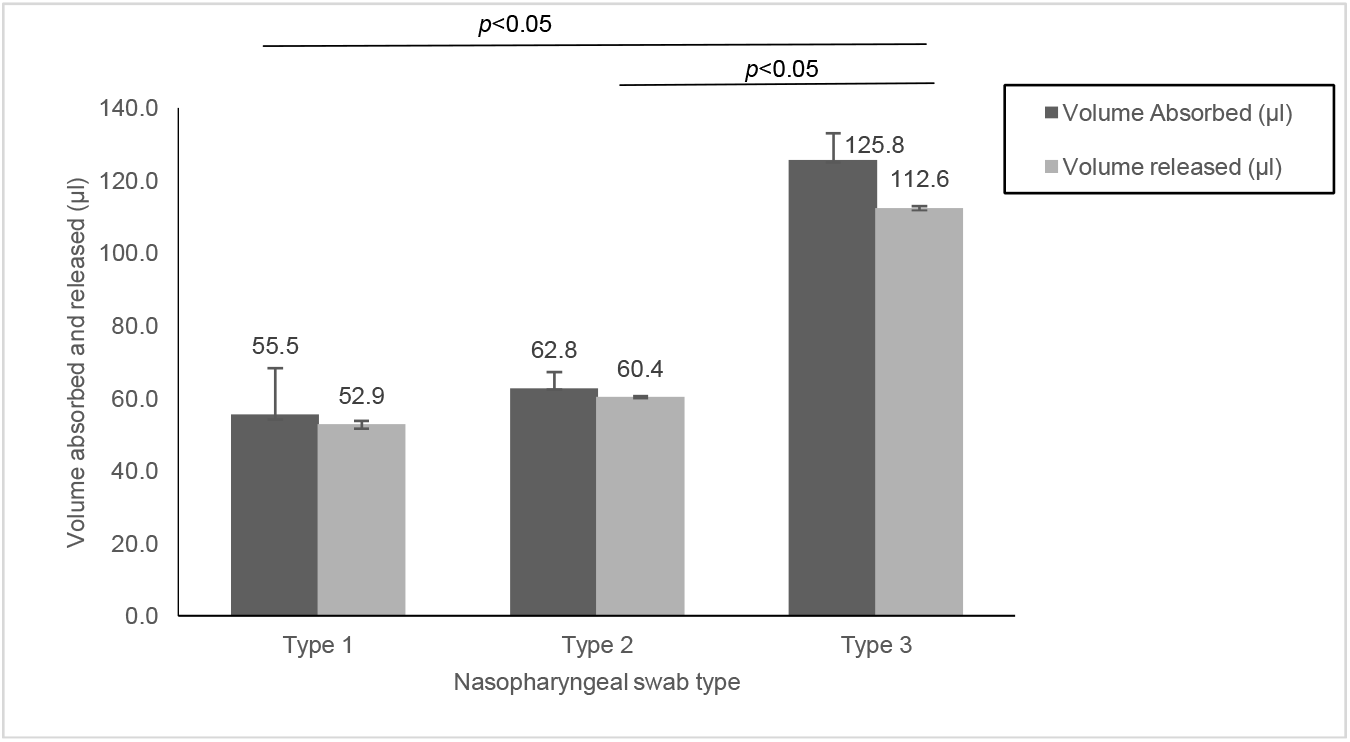
Volume of spiked PBS absorbed and released by the nasopharyngeal swabs. Data are presented as mean ± SEM. Type 1- South African supplier, Type 2- Kang Jian Medical Apparatus, China, Type 3- Wuxi NEST Biotechnology Co. Ltd, China.

### Swab viral capture, extraction and recovery efficiency analysis

Swab viral capture analysis showed an inverse relationship to volume absorbed. Type 3, the most absorbent swab had the least capture capabilities. However, the extraction and recovery efficiencies of each swab for SARS-CoV-2 RNA detection was strongly related to the type of swab and showed similar trends with the volume absorbed. We obtained the highest extraction and recovery efficiency for type 3 (12.4 – 38.9%) and the least extraction and recovery efficiency was obtained for type 1 (3.6 -24.7%) which shows a large part of the virus remained on the swab (Table 2).

**Table 1:** Qualitative characteristics of the different swab types

| Characteristic | Type 1 | Type 2 | Type 3 |
| --- | --- | --- | --- |
| Swab Material type-Texture | 5 | 4 | 4 |
| Swab Material type- Color | 5 | 4 | 5 |
| Shaft- material | 5 | 5 | 5 |
| Shaft properties | 5 | 5 | 4 |
| Shaft length | 4 | 5 | 5 |
| Breakpoint | 4 | 4 | 5 |
| Laboratory Ease of use | 5 | 5 | 5 |
| Robustness | 5 | 5 | 5 |
| Overall score | 38/40 | 37/40 | 38/40 |

The swab characteristics were differentiated using the 5-point Linkert scale of 1-5. Type 1- South African supplier, Type 2- Kang Jian Medical Apparatus, China, Type 3- Wuxi NEST Biotechnology Co. Ltd, China. Key: 1= very bad/difficult, 2= bad/difficult, 3= neutral, 4= good/easy, 5= very good/easy.

**Table 2:** viral capture, extraction and recovery efficiency analysis for the three NP swabs

| **Swab properties** | **Type 1** | **Type 2** | **Type 3** |
| --- | --- | --- | --- |
| Viral capture (%) | 29,6 | 24,4 | 22,2 |
| Extraction efficiency (EE) (%)day 0 | 5,2 | 6,3 | 13,1 |
| Extraction Efficiency (EE) (%)day 3 | 3,6 | 4,4 | 12,4 |
| Recovery efficiency (RE) (%) day 0 | 35,2 | 37,8 | 38,9 |
| Recovery efficiency (RE) (%) day 3 | 24,7 | 26,0 | 36,9 |

### Swab Testing: real-time RT-PCR

SARS-CoV-2 was detected in 100% of the NP swab types for all the viral culture dilutions and time points investigated. Generally lower Ct values were observed for swab types 2 and 3 at day 0 for the nucleocapsid 1 (*N1*) and nucleocapsid 2 (*N2*) genes at the lowest viral culture dilution (1:10 copies/ml). Higher Ct values were observed at day 3 indicating lower viral loads, suggesting some loss of stability of RNA on the swabs at day 3 (72 hours). We observed changes in Ct values for day 0 and day 3 across all swab types and viral dilutions, however, the differences in Ct values were not statistically significant (*N1* day 0: *p*= 0.9970; *N2* day 0: *p*=0.9984; *N1* day 3: *p*=0.8131; *N2* day 3: *p*= 0.6867) (Figure 3). No PCR inhibition or interference was observed for all the swab types and the negative and positive controls were valid.

**Figure 3:**
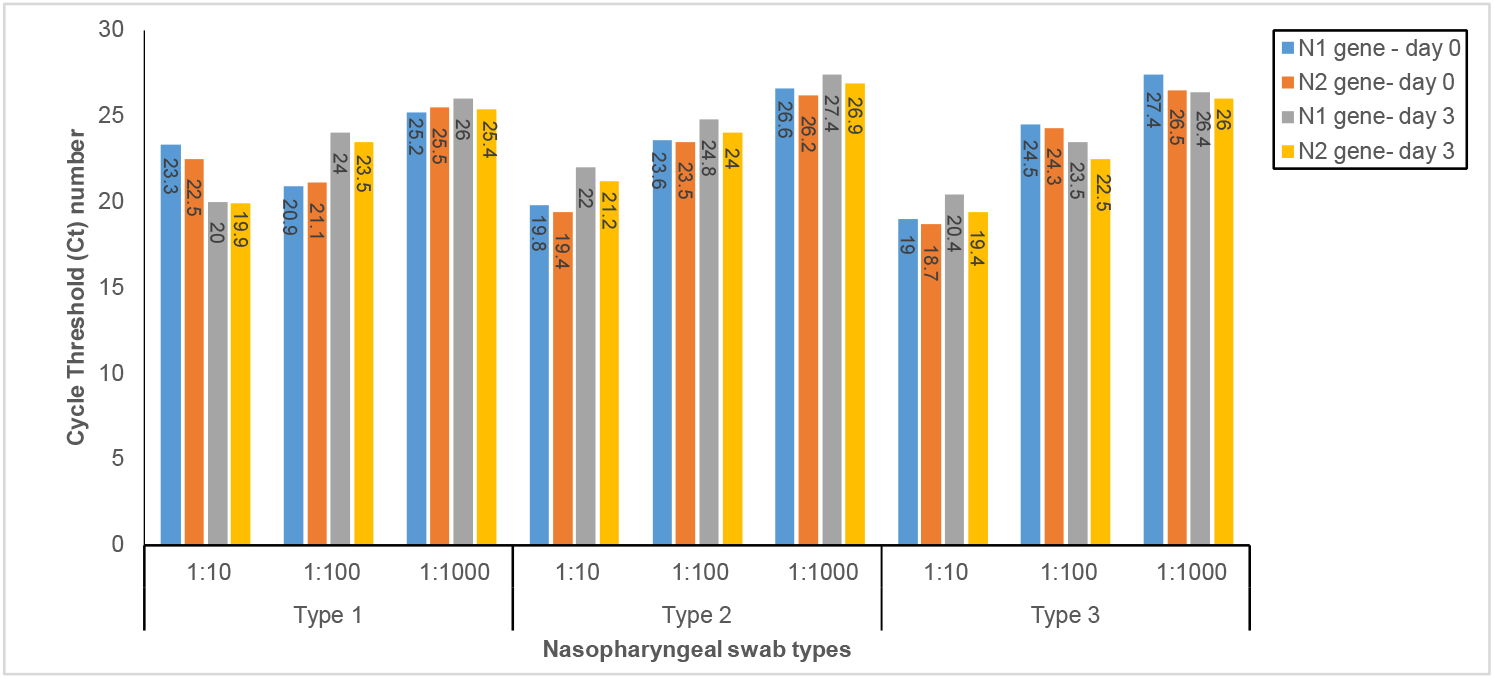
Detection of SARS-CoV-2 RNA by real-time RT-PCR analysis from nasopharyngeal swabs at day 0 and day 3. All viral culture dilutions were copies/ml. Type 1-South African supplier, Type 2-Kang Jian Medical Apparatus, China, Type 3-Wuxi NEST Biotechnology Co. Ltd, China.

## Discussion

Although several specimen types are listed as acceptable for SARS-CoV-2 diagnosis by the CDC and WHO, the NP swabs are still the gold standard method of sampling (4, 5, 20). These swabs are minimally invasive but discomfort during sampling has been highlighted in some studies inciting bleeding, nausea and vomiting in patients probably caused by the swab surface properties (9, 21, 22). In order to achieve maximal sample collection, the design and mechanical properties of the NP swabs is crucial. The recommended shaft length is ∼15cm and diameter of tip is ∼1-3.2mm. Other important properties include smoothness/softness, flexibility, ability to withstand torsion force, breakpoints, durability, collection sufficiency and PCR compatibility (21, 23). In this study we performed a qualitative analysis of the properties of three commercially available NP swabs. We observed some variations in the length of swab tip, shaft and breakpoints which ranged from 16-27mm, 150-152mm and 70-80mm respectively. All swabs had soft tips, which according to Copan Diagnostics facilitate efficient sample release (24). The colour of the swab tip has also been highlighted to be an important feature where generally a white swab tip indicates the absence of impurities/chemical coating/treatment of the swab, hence will aid in pure sample collection (25, 26). In the current study, this was observed for swab types 1 and 3.

The swab composition and structure have a significant impact on the main properties of the swab i.e. absorption, capture, extraction and recovery efficiency of collected sample (13, 14, 27). In our study, the three NP swabs revealed significant differences in volume absorption (the amount of fluid sample absorbed) and volume released (volume of fluid sample released into buffer). These parameters were inversely related to swab viral capture capacity (the amount of organisms removed from solution by the swab) but strongly related to extraction efficiency (the effectiveness of sample transfer from the swab tip to the extraction buffer) and recovery efficiency (the overall transfer effectiveness of swab from the sampling site to the extraction buffer). Type 3 NP swabs had the highest absorbing and releasing capacity compared to types 1 and 2 suggesting a direct relationship between the swab tip length (i.e. high amount of nylon flock material on the tip (refer to supplementary table 2)) and these properties. Less than 25%, 30% and 55% SARS-CoV-2 RNA was recoverable from swab types 1, 2 and 3 respectively, indicating that most of the viral particles either were left in buffer or were entrapped by the swab material.

The quality of swab samples and other factors such as sampling procedures, time of sampling and sample storage temperature contribute to adequate disease detection and monitoring of respiratory infections (12, 28, 29). We investigated storage of swabs over two time points (day 0 and day 3) at room temperature. RNA was detected from all swabs at both time points without PCR inhibition/interference but an increase in Ct values was observed at day 3 for all swab types, indicating lower viral loads detected. RNA stability was comparable for all swab types.

Although there is a vast amount of NP swab validation studies (2, 6, 14, 27, 30-34), this is, to our knowledge the first head-to-head comparison of the NP swab types from different manufacturers for SARS-CoV-2 testing. The limitations of this study include the unavailability of clinical specimens for comparison, broader storage times to investigate RNA stability on the swabs as well as inclusion of swabs from well-known manufacturer i.e. Copan and Puritan, which we could not acquire at the time of the study due to high-demand. Future focus would be to validate swabs in clinical settings on a larger population.

## Conclusion

We conclude that different nasopharyngeal swabs have different absorption, capture capabilities and extraction efficiencies. We recommend critical assessment of the swab features and properties before implementation for use at clinical settings in order to facilitate accurate diagnosis.

## Data Availability

Data is available

## Acknowledgements

We would like to thank Asiashu Singh-Moodley for assisting with critical reviews and staff at the Centre of Excellence for Biomedical TB Research at the University of the Witwatersrand (Bavesh Kana, Bhavna Gordhan, Edith Machowski, Wolfgang Preiser and Taejon Suliman) for assisting with viral cultures and mimetics. We also thank the Bill Melinda Gates Foundation for providing research funding support (grant no.OPP1171455).The funders had no role in study design, data collection and interpretation, or the decision to submit the work for publication.

---

**Supplementary Table S1:**
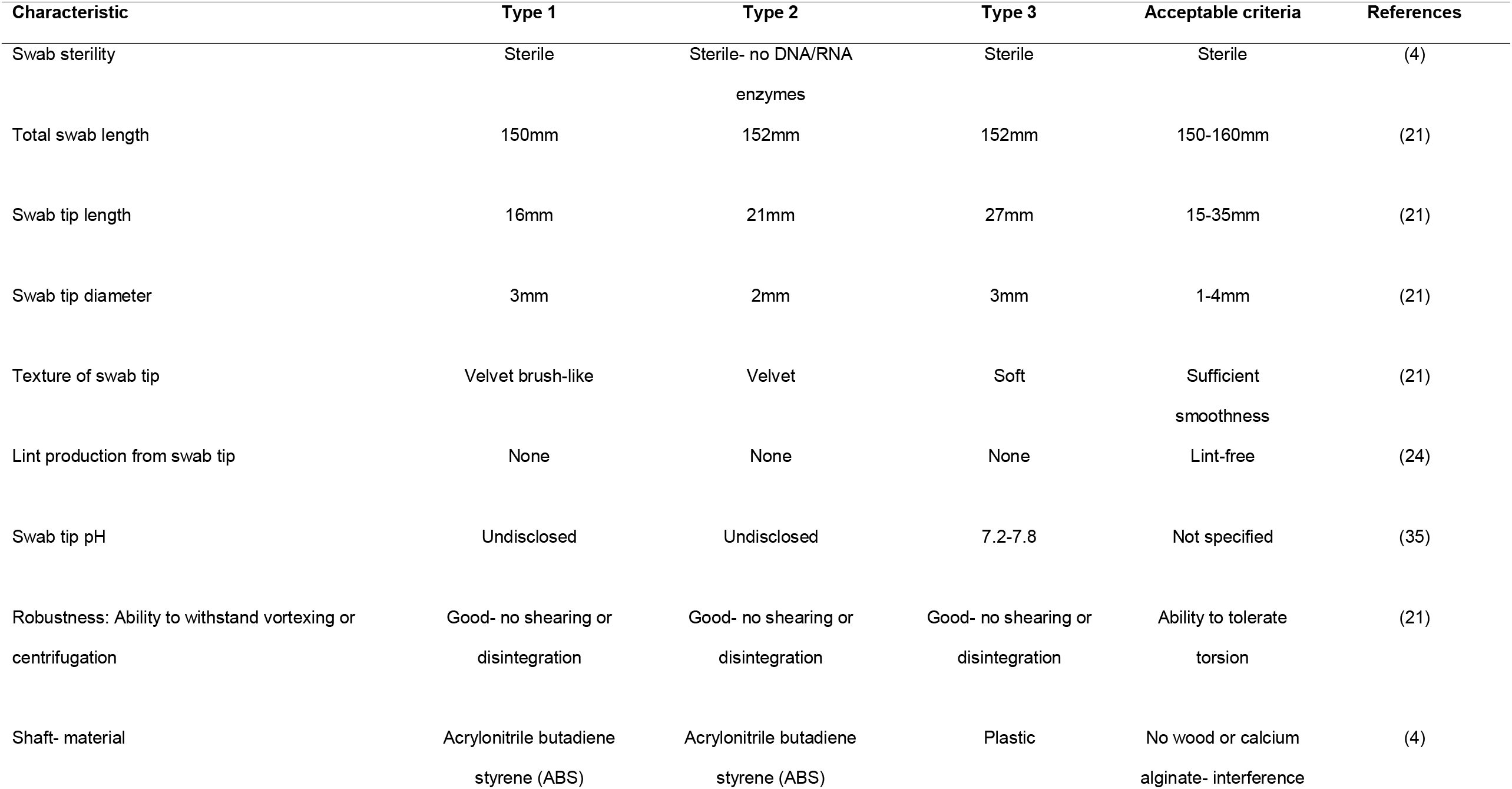
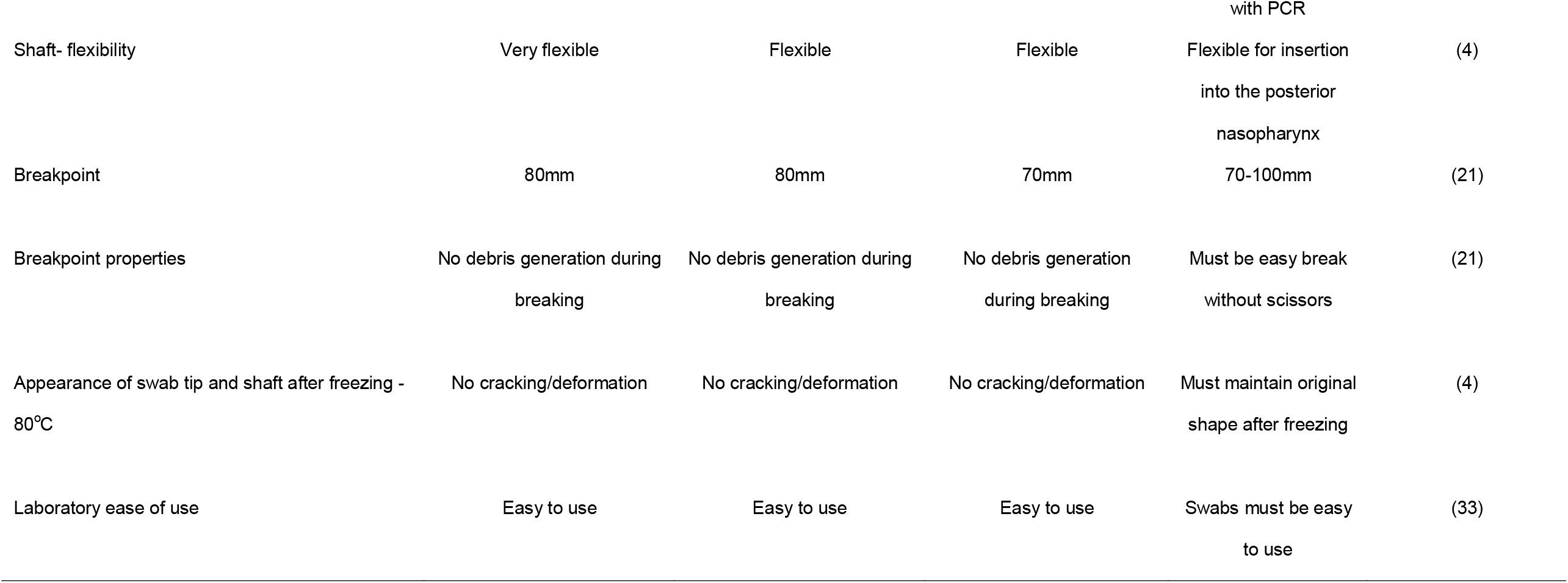
Characteristics of swabs and acceptance criteria

